# Cost-Performance Evaluation of Large Language Models for Aspect-Based Sentiment Analysis of HCAHPS Patient Comments: A Validation Study

**DOI:** 10.64898/2026.06.11.26355494

**Authors:** Khalid Nawab, Gretchen Ramsey, Samina Asfandiyar, Sayuj Atreya, Shadi Hijjawi, Sharatkumar Rokkam, Usman Ghayur, Akarshana Rajesh, Ihtesham Yousuf, Zefaf Ali Shah, Amit Kumar Misra, Madhushan Ponnala, Tauseef Hamid, Richard Schreiber

## Abstract

**Background:** Hospital Consumer Assessment of Healthcare Providers and Systems (HCAHPS) free-text comments contain actionable feedback, but timely, scalable, and affordable sentiment analysis remains challenging for health systems that rely on third-party vendors.

**Objectives:** To evaluate cost-performance tradeoffs between a cost-optimized and a flagship large language model (LLM) for aspect-based sentiment analysis of HCAHPS comments, using human inter-rater agreement as a reproducibility benchmark.

**Methods:** We analyzed 512 free-text HCAHPS comments collected from two community hospitals in calendar year 2023. Six trained reviewers (medical students, recent medical graduates, and practicing internists) independently assigned positive, negative, or neutral labels to each comment-aspect pair; the majority label among three reviewers formed the consensus reference standard. Two OpenAI models — GPT-5-nano (cost-optimized) and GPT-5 (flagship) — were prompted in a zero-shot setting via the OpenAI API. We calculated pairwise Cohen’s κ to establish a human inter-rater baseline, then compared each model’s labels to the consensus using Cohen’s κ, accuracy, weighted F1, and per-call cost and latency.

**Results:** Mean human inter-rater agreement was κ = 0.79 (substantial). Both LLMs exceeded this baseline (cost-optimized κ = 0.85; flagship κ = 0.85) with nearly identical accuracy (0.92) and weighted F1 (0.93 vs. 0.93). Performance was strong on positive (F1 ≈ 0.97) and negative (F1 ≈ 0.90) classes but poor on the underrepresented neutral class (F1 ≤ 0.19). The cost-optimized model processed all 512 comments for $0.04 versus $0.18 for the flagship — a 4.2-fold cost difference without measurable performance gain.

**Conclusions:** Commercially available LLMs can perform aspect-based sentiment analysis on HCAHPS comments at human-level reproducibility, with the cost-optimized tier sufficient for routine classification. This offers health systems a rapid, scalable, low-cost alternative to vendor-based patient-experience analytics.

## 1. Background and Significance

### 1.1. Financial Stakes of Patient Experience

The Hospital Value-Based Purchasing (VBP) Program is a budget-neutral initiative created under the Affordable Care Act. The Centers for Medicare and Medicaid Services (CMS) funds this program by withholding 2% of the base Diagnosis Related Group payments to the participating hospitals.^1^ This amount comes out to nearly $2 billion annually and is redistributed back to hospitals based on their Total Performance Score (TPS).^2^ Four domains equally determine a hospital’s TPS. Patient Experience of Care is one of them (along with Safety, Efficiency/Cost Reduction, and Clinical Outcomes). This means 25% of this “at-risk” revenue directly ties to patients’ feedback, which could potentially be millions of dollars in annual revenue.^2^ Furthermore, market competition has pushed hospitals to invest heavily in improving patients’ perception of their care via various amenities. A study found that such amenities cost hospitals more compared to improvement in clinical care quality, but improved amenities can lead to higher hospital volume.^3^ Ronald Reagan UCLA Medical Center saw an improvement from 71% to 85% in the number of patients that will definitely recommend the hospital after spending $829 million on rebuilding private rooms, views, and introducing massage therapy.^3^ This level of expenditure shows a clear belief by health systems that hospitality drives patient satisfaction and then hospital volume.

### 1.2. Cost–Performance Tradeoffs in Analyzing Patient Comments

Given the financial importance of patient experience metrics, hospitals must not only collect patient feedback but also analyze it quickly, consistently, and at scale. While prior efforts have focused on improving survey administration and response rates, less attention has been paid to the operational question of how best to analyze free-text patient comments once they are collected. In particular, healthcare systems face a practical decision when adopting large language models for sentiment analysis: whether lower-cost, lightweight models are sufficient for routine aspect-level sentiment classification, or whether more expensive flagship models provide meaningful performance advantages that justify their cost. This study addresses that question by evaluating the cost-performance tradeoffs of different LLM tiers using human inter-rater agreement as a reproducibility benchmark.

### 1.3. The Patient-Experience Feedback Lag

Despite the high investment figures, there is a significant lag in obtaining insights into patient experience. HCAHPS surveys, which are the national standard of patient experience measurement, are conducted either via mail or over the phone and the process is usually initiated 48 hours to six weeks post-discharge.^4^ 75% of these mailed surveys are returned 44 days after discharge, with a median of 43 days, according to real-world data.^5^ This delay is important because surveys that are sent back within 10 days get an overall rating of 78%, but those that are sent back about 44 days later get a rating of 71%. Results are usually reported every three months, but by then, opportunities for service recovery may have been missed and the collected data may not reflect the current situation. Poor performance in the patient experience domain of the CMS Hospital VBP program can reduce the hospital’s total performance score and lead to financial penalties.^2^

### 1.4. The Vendor Tax

Most health systems will outsource HCAHPS administration, data collection, and sentiment analysis to an approved third-party commercial vendor. These contracts can amount to a significant financial burden for the institution, especially with declining response rates and traditional administration modes.^6^ According to the CMS-approved vendor directory, basic HCAHPS survey administration costs range from $995 to $3,200 annually for small rural hospitals, with larger facilities paying $3,000 to $10,000 or more depending on patient volume.^7,8^ Furthermore, the vendors may charge more for services like advanced Natural Language Processing (NLP) and other analyses of the feedback data, making real time analytics more costly.^9^ Meanwhile other channels of patient feedback like nurse-rounding logs, grievance reports, patient portal messages, and online reviews remain unanalyzed.

### 1.5. Why Large Language Models (LLM)?

Sentiment analysis via traditional NLP approaches requires significant domain specific training data, as well as ongoing maintenance as language patterns evolve.^10^ LLMs on the other hand, without retraining or with just a few examples, can outperform traditional models as found by one study in which ChatGPT in a zero-shot setting (i.e., without task-specific training or labeled examples) outperformed various widely used sentiment analysis tools on health-related survey data without requiring any annotated training examples.^11^ Pre-trained on diverse corpora, LLMs can perform aspect-based sentiment analysis without task-specific fine-tuning and with just a few training examples via zero-shot or few-shot prompting.^12^ In clinical NLP, for tasks with limited training examples or with class imbalance, GPT-4 has been proven to outperform various supervised classifiers trained on annotated datasets.^13^ Similarly, LLMs show strong performance in aspect-based sentiment analysis of patient feedback outperforming traditional deep learning approaches.^14^ Thus LLMs provide an opportunity to analyze patient feedback rapidly without the overhead cost of training and maintenance of traditional classification models.

The advent of LLMs has led researchers to study LLM-based sentiment analysis for HCAHPS data. Zhang et. al. came up with a two-stage framework that combines LLMs with interpretable classifiers for explainable ABSA. They achieved macro-F1 score of 0.816 on HCAHPS comments, which is better than RoBERTa’s score of 0.764.^15^ This study, however, did not look at the cost-performance tradeoffs between LLM tiers. We try to fill this gap by comparing a cost-optimized LLM to a flagship LLM using a much simpler approach and establishing human inter-rater agreement as the reproducibility benchmark.

## 2. Objectives

Building on our previous work, where we demonstrated the utility of deep learning in extracting meaningful information from patient experience feedback comments,^16^ we hypothesize that large-language models can deliver real-time, omnichannel sentiment analytics with human-level precision, at a much lower cost compared to third party vendors. We contrasted a cost-effective LLM (designated as “cost-optimized”) with a premium flagship LLM using a dataset of 512 patient comments evaluated by three human reviewers. Our objectives were to (1) determine human inter-rater agreement; (2) quantify each model’s performance in relation to the consensus; (3) compare costs and processing times between the two models.

## 3. Methods

### 3.1. Dataset Description

The original dataset included 645 free-text comments collected via the HCAHPS survey between 01/01/2023 and 12/31/2023 in a column labeled as “Comment”, provided by the department of guest relations at Penn State Health. These were from patients discharged from Penn State Holy Spirit Medical Center and Penn State Health Hampden Medical Center. This dataset also contained a column “Coded as”, containing all aspects of care to which the “Comment” applies. Most comments contained opinions about more than one aspect of care. For example, consider this comment:

> “Care was excellent. Nursing staff were excellent! Bathroom smelled like sewer gas.”

This comment contains opinions regarding more than one aspect, “Nurse/Nurse Aid/Nursing” and “Cleanliness of Hospital Environment”. The original dataset also contained various other columns with information about the patient unit and hospital. This data was not included in the analysis as it was unrelated to our sentiment labeling task.

### 3.2. Data Labeling by Humans

During this stage, we presented comments with their respective aspects to human volunteers with the request to assign one of the three labels (“Positive”, “Negative” or “Neutral”) representing the sentiment carried in the comment regarding the presented aspect. We utilized 6 volunteers, 2 of whom were medical students, 2 medical school recent graduates, and 2 practicing internal medicine physicians. Considering that the same comment might contain opinions regarding multiple aspects, we asked participants to focus on the opinion/sentiment of the presented aspect only. The goal was to present each comment to at least three human participants. To make sure the reviewers understood the comment they were reviewing, they had the option to skip any comment if they felt they did not understand it due to language ambiguity or grammatical/spelling mistakes.

Once labeling by human volunteers was complete, comments where two of the three reviewers had assigned the same label were selected. This resulted in 512 comments. These labelled comments included those where at least two reviewers agreed on the same label. We then assigned a consensus label to each comment, representing the human majority (by two out of three reviewers) “Agreement”.

The 133 excluded comments contained comments where either the three reviewers assigned three different labels, or if the comment was skipped by reviewers and was reviewed by fewer than three reviewers.

### 3.3. Data Labeling by LLMs

We selected two publicly available LLMs with access provided by OpenAI via application programming interfaces (APIs). These models were:

- GPT-5-nano: At the time of this analysis, this was the cheapest available and thus cost-optimized model.^17^
- GPT-5: At the time of this analysis, this was the “flagship” or the “smartest” available model.^18^

We passed each row of the derived dataset with 512 rows, containing the comment, aspect pairs, to both OpenAI models via API, with the prompt in figure 1. We recorded the label returned by the model, along with the time taken by the API call (starting at the time the call was made to the time a response was returned), the input and output token counts, and calculated price for that specific call.

**Figure 1.**
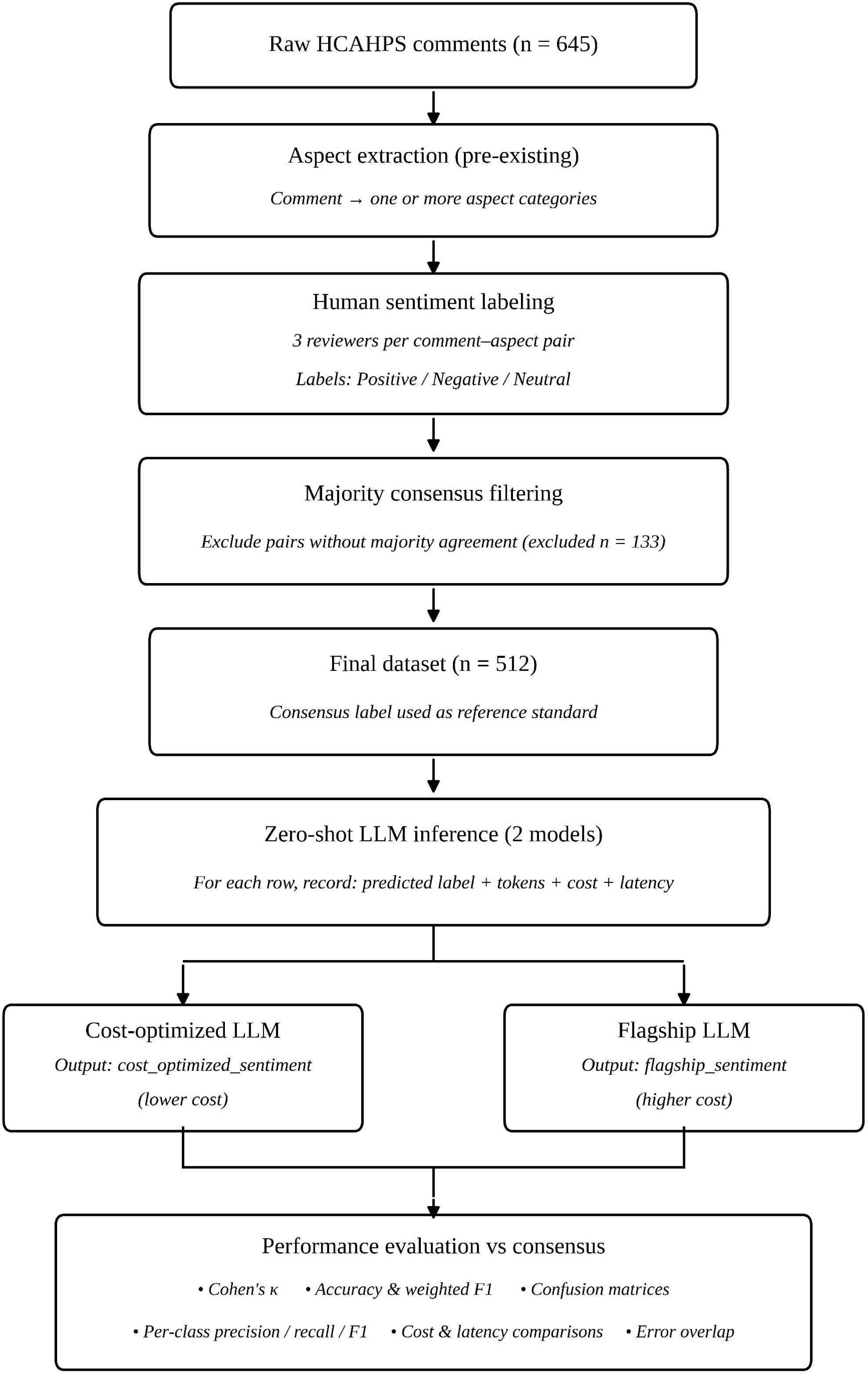
Zero-shot prompt template used for aspect-based sentiment classification of patient comments. The template specifies the comment text, the aspect under evaluation, and the three allowable sentiment labels (Positive, Negative, Neutral).

### 3.4. Statistical Evaluation

We conducted analysis in Python using publicly available data-science libraries pandas and scikit-learn.

#### 3.4.1. Measuring human inter-rater agreement and establishing human consensus

To quantify the level of agreement among human reviewers to establish the human reproducibility baseline, we calculated pairwise Cohen’s κ among the three reviewers. Cohen’s κ is commonly used to measure agreement beyond chance between 2 raters. Its values range from −1 to 1, with values between 0.61 and 0.80 commonly interpreted as substantial agreement.^19^ Since Cohen’s κ can only be applied to two raters, we computed it for reviewers in pairs (reviewer 1 vs 2, reviewer 1 vs 3 and reviewer 2 vs 3). We selected the mean of these three values to establish the human inter-rater agreement baseline. This baseline represents the reproducibility of human sentiment labeling and defines the level of agreement a model must achieve to be considered comparable to human performance.

For each comment, a single reference label (“consensus”) was constructed, which was derived from the label agreed upon by two out of three reviewers. This consensus label was then used as a reference standard for subsequent LLM evaluation.

#### 3.4.2. Measuring LLMs agreement with human consensus label

We computed Cohen’s κ between each model’s predicted labels and the human consensus labels. Each model’s κ value was interpreted relative to the human baseline rather than in isolation. A model achieving κ comparable to or exceeding the mean human inter-rater κ was considered to demonstrate human-level agreement.

#### 3.4.3. LLMs Performance Metrics

For each LLM, we calculated accuracy and weighted F1-score. Accuracy was defined as the proportion of comments for which the model’s predicted label matched the human consensus label. Weighted F1-score combines precision and recall, while weighting classes by their frequency, thus accounting for class imbalance among the three labels.

#### 3.4.4. Cost and Latency Analysis

For each model, the cost per API call was summed to calculate total cost and then divided by 512 to calculate mean cost per call. Similarly, processing time for all calls was summed and divided by 512 to calculate mean model latency time per call. Figure 2 shows a summary of our methodology.

**Figure 2.**
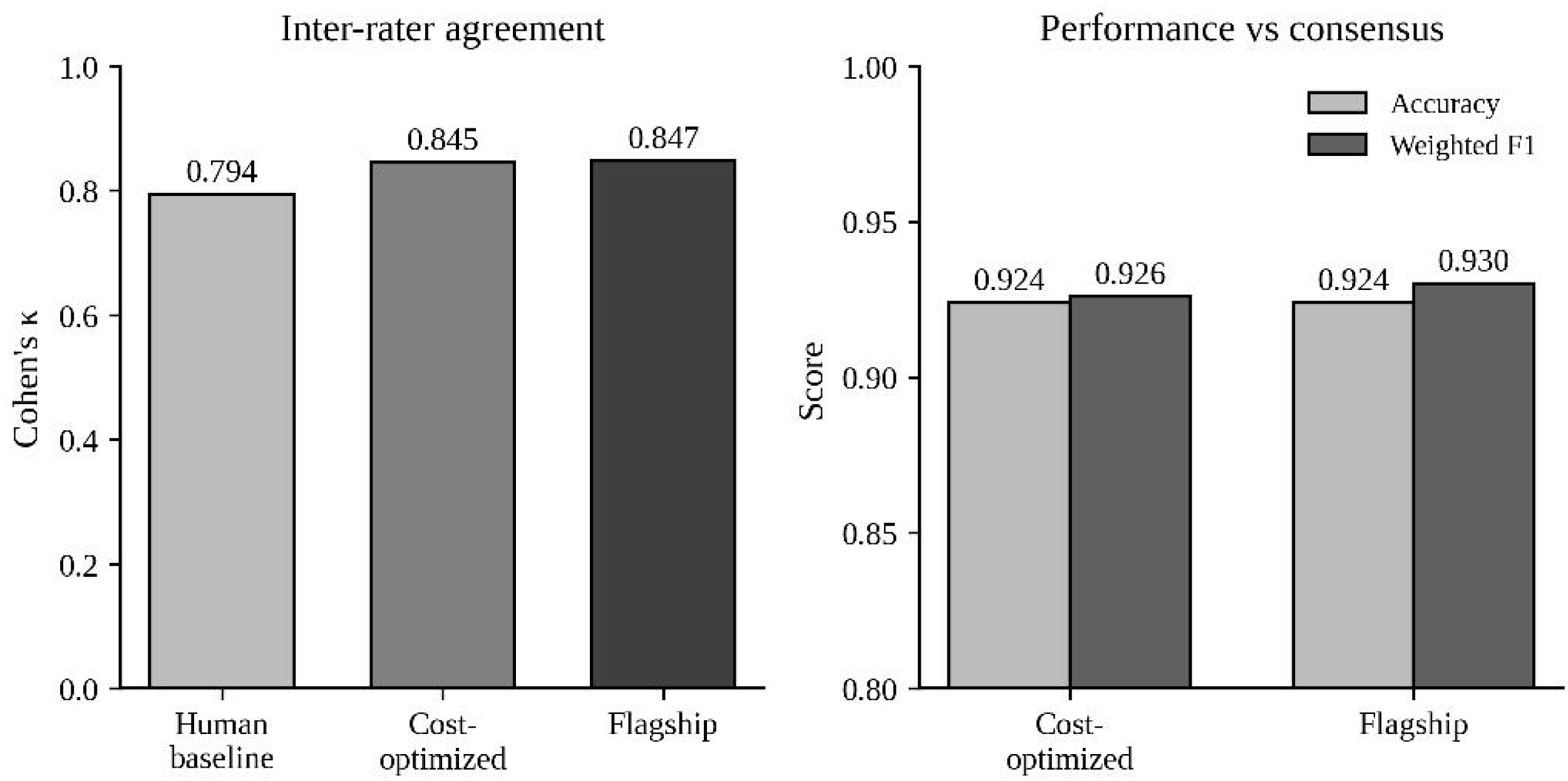
Study methodology overview. Workflow showing dataset derivation from 645 HCAHPS comments, three-reviewer human labeling, derivation of the 512-comment consensus subset, parallel zero-shot labeling by two LLMs (GPT-5-nano and GPT-5) via API, and downstream evaluation against the human consensus baseline.

### 3.5. Ethics Approval

This study was reviewed by the Pennsylvania State University Office for Research Protections, Human Research Protection Program (Study ID: STUDY00028653), and was determined to qualify as exempt research not requiring formal IRB review (Exemption Determination dated May 14, 2026). The analysis involved de-identified retrospective HCAHPS survey comments. The study was performed in compliance with the World Medical Association Declaration of Helsinki on Ethical Principles for Medical Research Involving Human Subjects.

## 4. Results

### 4.1. Label Distribution

Table 1 shows the distribution of consensus labels, highlighting substantial class imbalance, especially the scarcity of the neutral labels, which affects the interpretation of aggregate performance metrics.

**Table 1.** Distribution of the Consensus Labels.

| Label | n | % |
| --- | --- | --- |
| Positive | 315 | 61.5% |
| Negative | 186 | 36.3% |
| Neutral | 11 | 2.1% |

### 4.2. Human Baseline Agreement

Human inter-rater agreement as represented by pairwise Cohen’s κ ranged from 0.786 to 0.809, with a mean of 0.79, indicating substantial agreement. Thus, we concluded for any model to be considered to have human-level reproducibility, it must match this level of agreement with the human consensus.

### 4.3. Model Performance

Both LLMs achieved a significant level of agreement with the consensus with Cohen’s κ for both models very close to each other (0.84 and 0.85). Similarly, the accuracy and weighted-F1 scores were also essentially identical for both models. See table 2. Figure 3 shows that both LLMs did better than the human baseline in terms of inter-rater agreement and got similar accuracy and F1-scores.

**Figure 3.**
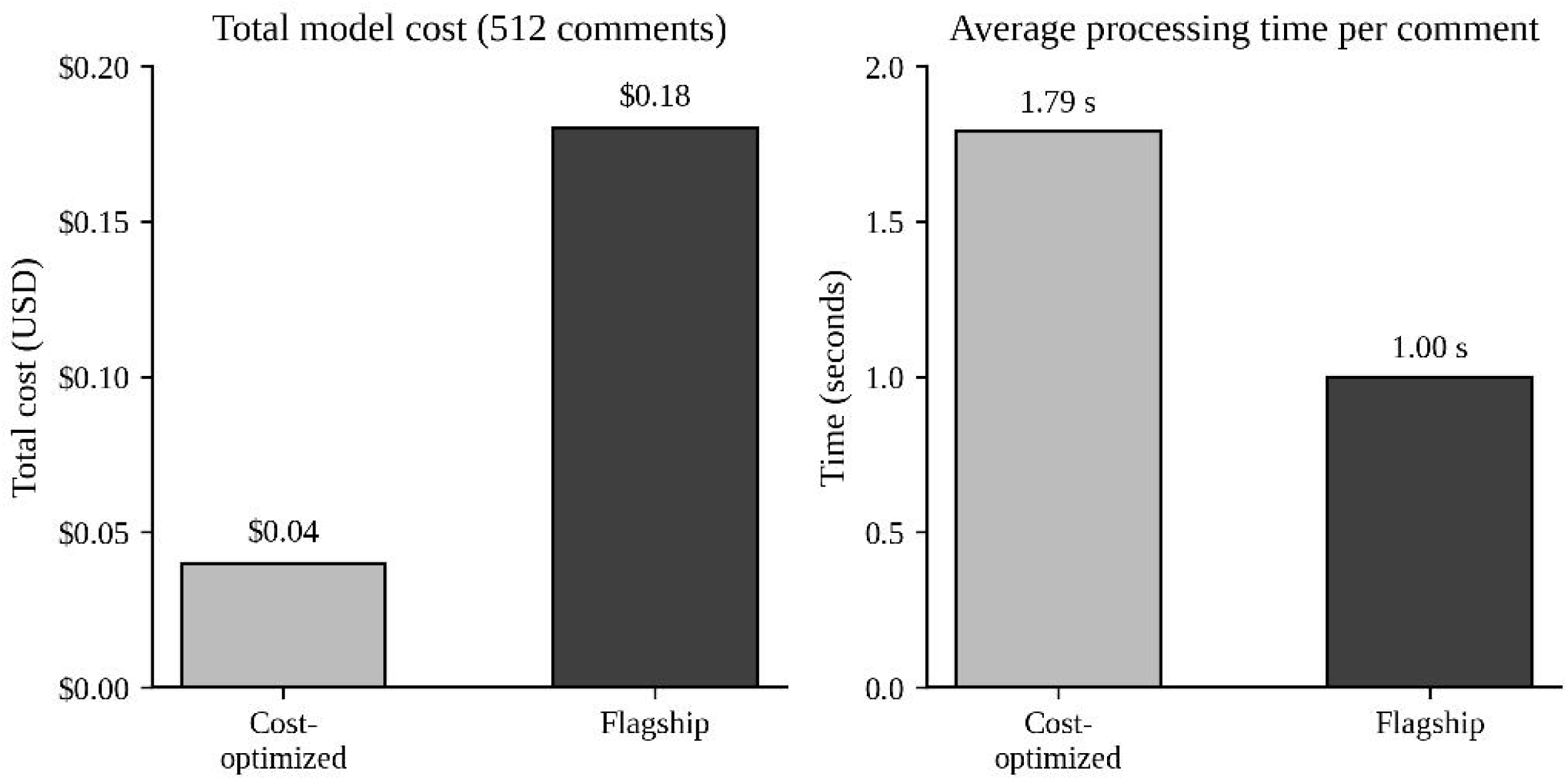
Inter-rater agreement and model performance. Both models achieved κ ≈ 0.85 against the reviewer consensus, exceeding the human baseline (κ ≈ 0.79). Accuracy and weighted F1 scores were similar for both models (approximately 0.93).

**Table 2.** Model Performance Metrics.

| Metric | Human Baseline | Cost-Optimized | Flagship |
| --- | --- | --- | --- |
| Cohen's $\kappa$ | 0.794 | 0.845 | 0.847 |
| Accuracy | — | 0.924 | 0.924 |
| Weighted F1 | — | 0.926 | 0.930 |
| Macro F1 | — | 0.676 | 0.690 |

### 4.4. Per-Class Performance

Performance of the LLMs varied significantly across the three classes as shown in Table 3. The majority of the comments as classified by the consensus label were labelled Positive, followed by Negative. Both models did very well on these majority classes, but performance was very poor for Neutral comments, highlighting the challenge of identifying ambivalent or mixed sentiment comments.

**Table 3.** Per-Class Performance (F1-Score).

| Class | n | Cost-Optimized F1 | Flagship F1 |
| --- | --- | --- | --- |
| Positive | 315 | 0.967 | 0.971 |
| Negative | 186 | 0.902 | 0.905 |
| Neutral | 11 | 0.160 | 0.194 |

### 4.5. Error Analysis

The error rate for both models was 7.6 % (39/512). Both models shared 31 errors out of the 39 (79.5%) where they disagreed with the human consensus. Interestingly, both models made errors on 8 comments where they disagree with the human consensus, but these were different comments for each model, indicating that each model makes partially independent errors despite similar aggregate performance.

### 4.6. Cost and Latency Analysis

The cost for processing 512 comments for each model can be considered minimal as it was less than $1. Figure 4 shows the difference in price: the cost-optimized model processed all 512 comments for about $0.04, while the flagship model cost $0.18, which is 4.2 times more without improved performance. The processing time for the cost-optimized model was a little longer (about 1.79 seconds per comment).

**Figure 4.**
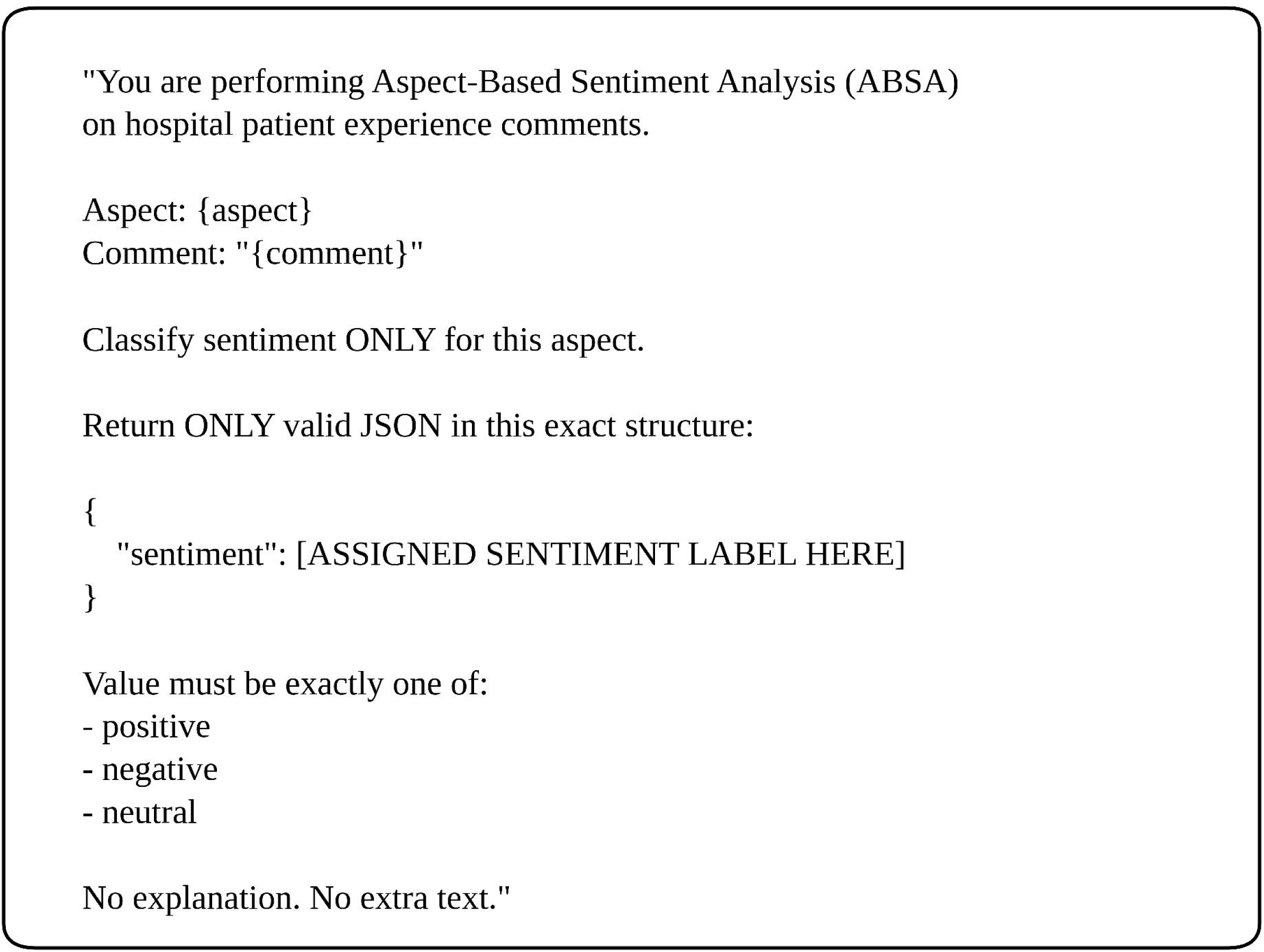
Cost and processing time comparison for both LLMs. The cost-optimized model (GPT-5-nano) processed all 512 comments for $0.04 with a mean latency of 1.79 seconds per comment; the flagship model (GPT-5) cost $0.18 — 4.2× more expensive — but was faster, at 1.00 seconds per comment on average.

## 5. Discussion

Our study demonstrates that commercially available LLMs accessible via APIs can perform aspect-based sentiment analysis on patient feedback comments at a performance level equal to human consensus very rapidly and at a cost orders of magnitude below commercial third-party vendors. This can enable healthcare systems to perform real-time analysis of patient feedback affordably and at scale.

Several of our findings are worth highlighting. The cost-optimized model performed as well as the flagship model at a lower cost. This suggests that for straightforward classification, one may not need to use more expensive models, and the cost-optimized models are enough. Even though the cost-optimized model is advertised as faster with low inference time by OpenAI, in our testing, it had a longer mean time taken per classification task. This is likely because the longer response time for the flagship model as advertised is for tasks requiring reasoning whereas sentiment classification task may be a relatively simple task for these models which they can perform quickly without much reasoning. Both models also exceeded human inter-rater agreement criterion standard. This indicates that classification tasks by LLMs are at least as reproducible as human judgment, a fact very relevant for operational deployment.

The error analysis showed that both LLM models failed to agree with human consensus over the same comments for the most part. This overlap between both the models suggests systemic difficulty with certain comment types rather than random error. This also suggests that using more expensive models may not translate into a proportionate performance benefit.

Our work adds to recent research by Zhang et.al.^15^ Their work prioritized explainability and interpretability. We addressed a different operational question: whether a cost-optimized LLM can match a flagship model in terms of performance for routine classification tasks. Furthermore, their framework required substantial implementation effort, adding to engineering overhead, whereas a zero-shot out-of-the-box approach implementing APIs offers a lower barrier to adoption for health systems seeking rapid deployment. To contextualize the cost implications, imagine a healthcare system has to process 10,000 HCAHPS comments. The processing cost at the price rate at the time of our study, using the cost-optimized LLM would be about $0.78 and using the flagship LLM would be about $3.48.

Our study has several strengths. It establishes a human inter-rater agreement as a reproducibility benchmark (κ=0.79) as a meaningful threshold for comparison with LLM performance instead of relying solely on accuracy metrics. While prior work focused on comparison of LLMs with traditional NLP methods, like we did in our previous work,^16^ our current work provides a direct cost and performance comparison of various LLM tiers, systemically answering the question of whether premium models justify their cost. We employed a zero-shot prompting approach without using any optimization and fine-tuning, prioritizing generalizability.

We recognize important limitations of our study, starting with severe class imbalance, with the comments classified as Neutral by human-consensus were only 2.1% of all the comments analyzed, with both models performing poorly on these comments. Thus, for practical use, enriched sampling or class specific prompting may have to be implemented. We also recognize that we lost about 20.6% of the initial dataset due to human reviewers not achieving majority consensus or reviewers choosing to skip some comments due to ambiguity. This raises concerns about selection bias. The retained sample may overrepresent simpler cases where it was easier to classify the sentiment. Future work should focus on examining model behavior on high-disagreement cases.

We also evaluated a simple prompt without optimization or sensitivity analysis. Prompt engineering can potentially improve performance, especially on edge cases or neutral comments, however, this would also reduce generalizability claimed by zero-shot approaches.

While our analysis used a limited sample of comments, these findings indicate potential for scalable, low-cost, real-time analytics, though system-level integration and infrastructure factors were not evaluated.

We only looked at models from one vendor (OpenAI), which makes it hard to apply the results to other LLM platforms. Anthropic (Claude), Google (Gemini), and Meta (Llama) all offer similar commercial LLMs. There are also open-source models like Mistral that can be used locally, which is important for health systems that are worried about sending protected health information to third-party APIs. Different providers may have different performance, cost, and latency characteristics, and pricing structures that are specific to each vendor change often. Our findings should be viewed as evidence of the viability of LLM-based HCAHPS comment analysis rather than as a validation of a particular platform. Future work should compare the performance of different vendors and investigate open-source options that can be deployed locally and may better meet the data governance needs of the institution.

## 6. Conclusions

Our study demonstrates that commercially available LLMs accessible via APIs can perform aspect-based sentiment analysis on patient feedback comments at a performance level comparable to human consensus, rapidly and at a cost order of magnitude below commercial third-party vendors. Additionally, on our proof-of-concept dataset, a cost-optimized LLM performed nearly identically to a flagship model, suggesting that lower-cost models may suffice for routine sentiment classification tasks. These models can be accessed conveniently via APIs at a minimal cost and without the need for deploying complex framework. Using the zero-shot approach, no model training is required. This provides a great opportunity for health systems that are looking to analyze patient feedback in real time.

## Clinical Relevance Statement

Health systems can deploy commercially available large language models via API to perform aspect-based sentiment analysis on HCAHPS and other patient-experience comments at human-level reproducibility and at a fraction of typical vendor cost. A cost-optimized model tier appears sufficient for routine classification, enabling near-real-time service-recovery workflows and broader analysis of free-text patient feedback channels that are currently unanalyzed.

## Data Availability

All data produced in the present study are available upon reasonable request to the authors.

## Acknowledgments

The authors thank the Department of Guest Relations at Penn State Health for providing access to the de-identified HCAHPS comment dataset used in this study.

## Protection of Human and Animal Subjects

The study was performed in compliance with the World Medical Association Declaration of Helsinki on Ethical Principles for Medical Research Involving Human Subjects. The study was reviewed by the Pennsylvania State University Office for Research Protections, Human Research Protection Program (Study ID: STUDY00028653) and determined to qualify as exempt research not requiring formal IRB review (Exemption Determination dated May 14, 2026). The analysis involved de-identified retrospective HCAHPS survey comments. No animal subjects were involved.

## References

1. Hospital Value-Based Purchasing | CMS. Accessed April 4, 2026. https://www.cms.gov/medicare/quality/value-based-programs/hospital-purchasing

2. Value-Based Purchasing: What Every Hospital Leader Must Know - Ambient. Accessed December 7, 2025. https://ambientclinical.com/2025/08/26/value-based-purchasing-what-every-hospital-leader-must-know/

3. Goldman DP, Vaiana M, Romley JA. The Emerging Importance of Patient Amenities in Hospital Care. N Engl J Med 2010;363(23):2185. doi:10.1056/NEJMP1009501

4. CMS. The HCAHPS Survey-Frequently Asked Questions What items are on the HCAHPS Survey? Accessed December 7, 2025. https://www.qualitynet.org/inpatient/hvbp

5. Moving the HCAHPS Needle - NRC Health. Accessed December 19, 2025. https://nrchealth.com/resource/full-report-moving-the-hcahps-needle/

6. Bland C, Zuckerbraun S, Lines LM, et al. Challenges Facing CAHPS Surveys and Opportunities for Modernization. RTI Press Published online November 16, 2022. doi:10.3768/RTIPRESS.2022.OP.0080.2211

7. National Rural Health Resource Center. Hospital Consumer Assessment of Healthcare Providers and Systems (HCAHPS) Overview Vendor Directory. February 2018. Accessed January 13, 2026. https://ruralhealth.utah.gov/wp-content/uploads/QR_HCAHPS-Vendor-Directory-2019-08.pdf

8. HCAHPS Online. Approved Vendor List: HCAHPS Approved Survey Vendors as of November 13, 2025. November 13, 2025. Accessed January 13, 2026. https://hcahpsonline.org/en/approved-vendor-list/

9. Assessing the Market for Patient Experience Surveying | TechTarget. Accessed December 26, 2025. https://www.techtarget.com/patientengagement/news/366584968/Assessing-the-Market-for-Patient-Experience-Surveying

10. Villanueva-Miranda I, Xie Y, Xiao G. Sentiment analysis in public health: a systematic review of the current state, challenges, and future directions. Front Public Health 2025;13. doi:10.3389/FPUBH.2025.1609749

11. Lossio-Ventura JA, Weger R, Lee AY, et al. A Comparison of ChatGPT and Fine-Tuned Open Pre-Trained Transformers (OPT) Against Widely Used Sentiment Analysis Tools: Sentiment Analysis of COVID-19 Survey Data. JMIR Ment Health 2024;11(1). doi:10.2196/50150

12. Zhang W, Deng Y, Liu B, Pan SJ, Bing L. Sentiment Analysis in the Era of Large Language Models: A Reality Check. Findings of the Association for Computational Linguistics: NAACL 2024 - Findings Published online May 24, 2023:3881–3906. doi:10.18653/v1/2024.findings-naacl.246

13. Sushil M, Zack T, Mandair D, et al. A comparative study of large language model-based zero-shot inference and task-specific supervised classification of breast cancer pathology reports. J Am Med Inform Assoc 2024;31(10):2315–2327. doi:10.1093/JAMIA/OCAE146

14. Alkhnbashi OS, Mohammad R, Hammoudeh M. Aspect-Based Sentiment Analysis of Patient Feedback Using Large Language Models. Big Data and Cognitive Computing 2024, Vol 8, Page 167 2024;8(12):167. doi:10.3390/BDCC8120167

15. Zhang Y, Wen S, Zhu Y, Li Z, Wang X. Combining large language models with interpretable models for explainable aspect-based sentiment analysis in the medical domain. Journal of King Saud University Computer and Information Sciences 2025 37:7 2025;37(7):175. doi:10.1007/S44443-025-00194-0

16. Nawab K, Ramsey G, Schreiber R. Natural Language Processing to Extract Meaningful Information from Patient Experience Feedback. Appl Clin Inform 2020;11(2):242. doi:10.1055/S-0040-1708049

17. GPT-5 nano Model | OpenAI API. Accessed December 19, 2025. https://platform.openai.com/docs/models/gpt-5-nano

18. GPT-5 Model | OpenAI API. Accessed December 19, 2025. https://platform.openai.com/docs/models/gpt-5

19. Landis JR, Koch GG. The measurement of observer agreement for categorical data. Biometrics 1977;33(1):159–174

